# Characterization of copy number alterations in Pakistani patients with colorectal cancer

**DOI:** 10.1101/2023.06.19.23291601

**Authors:** Clayton T. Marcinak, Bradon R. McDonald, Hina Zubairi, Sabeehuddin Siddique, Michelle D. Stephens, Munira Moosajee, Rashida Ahmed, Syed Nabeel Zafar, Muhammed Murtaza, Kulsoom Ghias

## Abstract

**Purpose:** Incidence and mortality rates of colorectal cancer (CRC) in South Asia are expected to rise by 70% between 2020 and 2040. Despite recent advances in the understanding of molecular alterations in CRC, the representation of South Asians in published genomic datasets is very limited. To address this gap, we performed a pilot study based on a transnational collaboration between academic centers in the US and Pakistan to characterize copy number alterations in a Pakistani cohort with CRC.

**Methods:** We obtained archived formalin-fixed paraffin-embedded (FFPE) tissue samples from 43 CRC patients treated at Aga Khan University (AKU) Hospital. DNA was extracted and evaluated for quality at AKU, and whole genome sequencing library preparation and shallow whole genome sequencing was performed at University of Wisconsin–Madison. Sequencing data was aligned to the human genome, and we used ichorCNA and GISTIC2 to determine relative copy number alterations.

**Results:** After removing 13 samples with insufficient DNA quality or quantity, tumor sequencing data was analyzed from 30 patients. The cohort consisted of 19 (63.3%) males with a mean age of 50.1 years and standard deviation (SD) of 17.8 years. Twenty-one (70.0%) patients had tumor stage T3 or T4, and 15 (50.0%) had tumors of the colon. A mean of 265 ng (SD, 29 ng) of DNA was extracted from each FFPE tumor sample, and each sample was sequenced to a mean depth of 0.22× (0.21×). Analysis of copy number alterations revealed recurrent gains in regions 8q and 13q, as well as losses in region 8p.

**Conclusions:** This transnational pilot study helped identify the challenges related to FFPE sample quality that can impede similar efforts in the future. Our results revealed similarities in the most common copy number alterations in CRC identified in native Pakistani patients and published datasets.

## Introduction

Colorectal cancer (CRC) is the third most common cancer worldwide and is responsible for nearly two million cases and one million deaths each year.^1^ In Pakistan, CRC ranked in the top ten cancers in terms of both incidence and mortality in the year 2020.^2^ Furthermore, low- and middle-income countries (LMICs) are expected to demonstrate a steep increase in cancer incidence and mortality due to changes in diet and environmental exposures.^3^

Since the founding of The Cancer Genome Atlas (TCGA) project in 2005, considerable investment has been made to characterize the molecular alterations underlying many cancer types, including CRC.^4^ As a result, investigators and clinicians alike have gained valuable insight into disease-causing and potentially targetable genomic changes in CRC, including mutations in *APC* and *KRAS* and copy number alterations in *MYC* and *ERBB2*.^4,5^ However, TCGA draws overwhelmingly from patients of European descent. A 2020 analysis by Carrot-Zhang *et al*. examined 10,678 individuals from TCGA, of whom over 80% were of European descent.^6^ While South Asians constitute approximately 25% of the world’s population, only 0.25% of participants in the TCGA—27 patients total—were of South Asian descent. Genomic characterization data of cancer in South Asian populations is very limited, and whether these tumors have similar somatic genomic alterations compared to those described in TCGA is unknown. This knowledge gap has multiple downstream implications, including the inability to design clinical trials guided by actionable cancer mutations, the inability for clinicians to evaluate and apply new advances in therapeutics and diagnostics for South Asian patients, and missed opportunities to further understand carcinogenesis in distinct genetic and environmental backgrounds.

There are several barriers that drive under-representation of patients from LMICs in global efforts to characterize cancer genomes. These barriers include limited research funding, minimal access to expensive cutting-edge genomic technology, fewer personnel with the required skillset for genomic analysis, and a lack of genomics-friendly sample processing workflows. Recent advances in sequencing technologies have significantly lowered the upfront investment and recurring costs for sequencing data generation, yet there is still a significant need to improve local computational and technical know-how in LMICs. One way to address this knowledge gap is to establish transnational collaborations that leverage high-throughput genomics technologies widely available in academic centers across the United States to characterize tumor and blood samples from patients with cancer living in LMICs. These collaborations can create opportunities for capacity building, skills training in genomics and precision medicine, and identifying promising avenues for further research.

Here, we report results from one such pilot collaboration, in which we harness efficient and scalable shallow whole genome sequencing (WGS) to broaden our limited understanding of genomic changes driving colorectal cancer carcinogenesis in the Pakistani population.

## Methods

### Study design and informed consent

This study was performed following review and approval by the Ethical Review Committee at Aga Khan University (Reference #: 0442; originally 4665-BBS-ERC-17) and Pakistan’s National Bioethics Committee (Reference #: 4-87/NBC-251). We identified available formalin-fixed paraffin-embedded (FFPE) tumor blocks from surgical resections performed for CRC between 2009 and 2014 in the archives of the Department of Pathology and Laboratory Medicine at Aga Khan University Hospital (AKUH) in Karachi, Pakistan. The histopathology service at AKUH receives tissue samples from surgical resections performed at the hospital, as well as from referring hospitals within the country. To include these samples in the current study, we contacted the patients or their next of kin to obtain informed consent for use of samples.

### DNA extraction

We stained mounted tissue sections using hematoxylin and eosin (H&E) to identify tumor regions within each FFPE tissue block. Tumor cellularity of patient material in each block was determined by histopathologists. We isolated DNA from identified tumor sections of each block into 20 μL of suspension buffer using the QIAmp DNA FFPE Tissue Kit as per manufacturer’s protocol (QIAGEN, Hilden, Germany). Quality and quantity of isolated DNA was determined using NanoDrop spectrophotometry and Qubit fluorometry (both from ThermoFisher Scientific, Waltham, MA, USA). After extraction and quality assessment at AKUH, DNA samples were shipped to the University of Wisconsin–Madison (UW–Madison) for library preparation and sequencing.

### Whole genome sequencing

We sheared the DNA by sonication using the Bioruptor Pico (Diagenode Inc., Denville, NJ, USA) as needed to achieve a target template size of 300 bp. Shearing settings varied from 0 to 6 cycles of one minute duration (30 seconds on, 30 seconds off), depending on initial assessment of fragment size distribution evaluated using the 4200 TapeStation System (Agilent Technologies, Santa Clara, CA, USA). We prepared WGS libraries from the extracted DNA using the Takara ThruPLEX HV kit (Takara Bio USA, San Jose, CA, USA), as per manufacturer’s protocol. Sequencing libraries were assessed for quality and concentration using the High Sensitivity D1000 Assay on 4200 TapeStation. We generated sequencing data using the NextSeq 1000 System (Illumina Inc., San Diego, CA, USA), generating 100 bp paired-end reads. After on-board FASTQ conversion and sample demultiplexing, sequencing data were aligned to human genome build hg19 using BWA-MEM.^7,8^ We marked duplicate reads and calculated alignment statistics using SAMtools.^9^

### Analysis of copy number aberrations

We used ichorCNA version 0.3.2 to perform copy number analysis of all autosomal chromosomes with 500kb windows.^10^ ichorCNA evaluates the relative ratio in each 500kb bin between a test sample and a panel of normal DNA samples. Based on changes in these ratios across the genome, these bins are grouped in segments and annotated for copy number aberrations. Prior to ichorCNA analysis, we calculated read depth at each bin using BEDTools.^11^ To establish baseline variation in read depth for ichorCNA using normal DNA samples, we used shallow WGS data generated from DNA from a panel of 16 adjacent normal FFPE tissue. These adjacent normal samples were also obtained from the histopathology archive at AKUH and evaluated to be tumor negative by histopathology. Normalized log2 depths for each bin calculated by ichorCNA were used as the input for Genomic Identification of Significant Targets in Cancer, version 2.0 (GISTIC2), to evaluate whether any observed copy number alterations were statistically enriched in this cohort.^12^

### Statistical analysis

Descriptive statistics were performed using frequency for categorical variables and mean with standard deviation (SD) for continuous variables. Tumor cellularity is presented as median and interquartile range (IQR). All analyses were performed using Julia, version 1.7,^13^ and R, version 4.2.2 (R Project for Statistical Computing, Vienna, Austria; www.r-project.org).

## Results

### Cohort characteristics

Tumor blocks from 43 patients underwent DNA extraction and quality assessment. Of these, 13 samples were deemed ineligible for analysis due to either extensive DNA degradation or insufficient DNA quantity. The final cohort consisted of 30 patients: 19 (63.3%) males and 11 (36.7%) females, with an average age of 50.1 years (SD, 17.8 years) at the time of diagnosis.

Age of diagnosis was unknown for two patients (6.7%). The majority of the cohort (21 patients, 70.0%) had T3 or T4 tumors at the time of histopathologic examination, while 16 patients (53.3%) had node-negative disease. Fifteen patients (50.0%) had a tumor of the colon, while 12 (40.0%) had more distal tumors of the rectosigmoid junction or rectum. The complete clinical characteristics of the cohort are shown in Table 1.

**Table 1:**
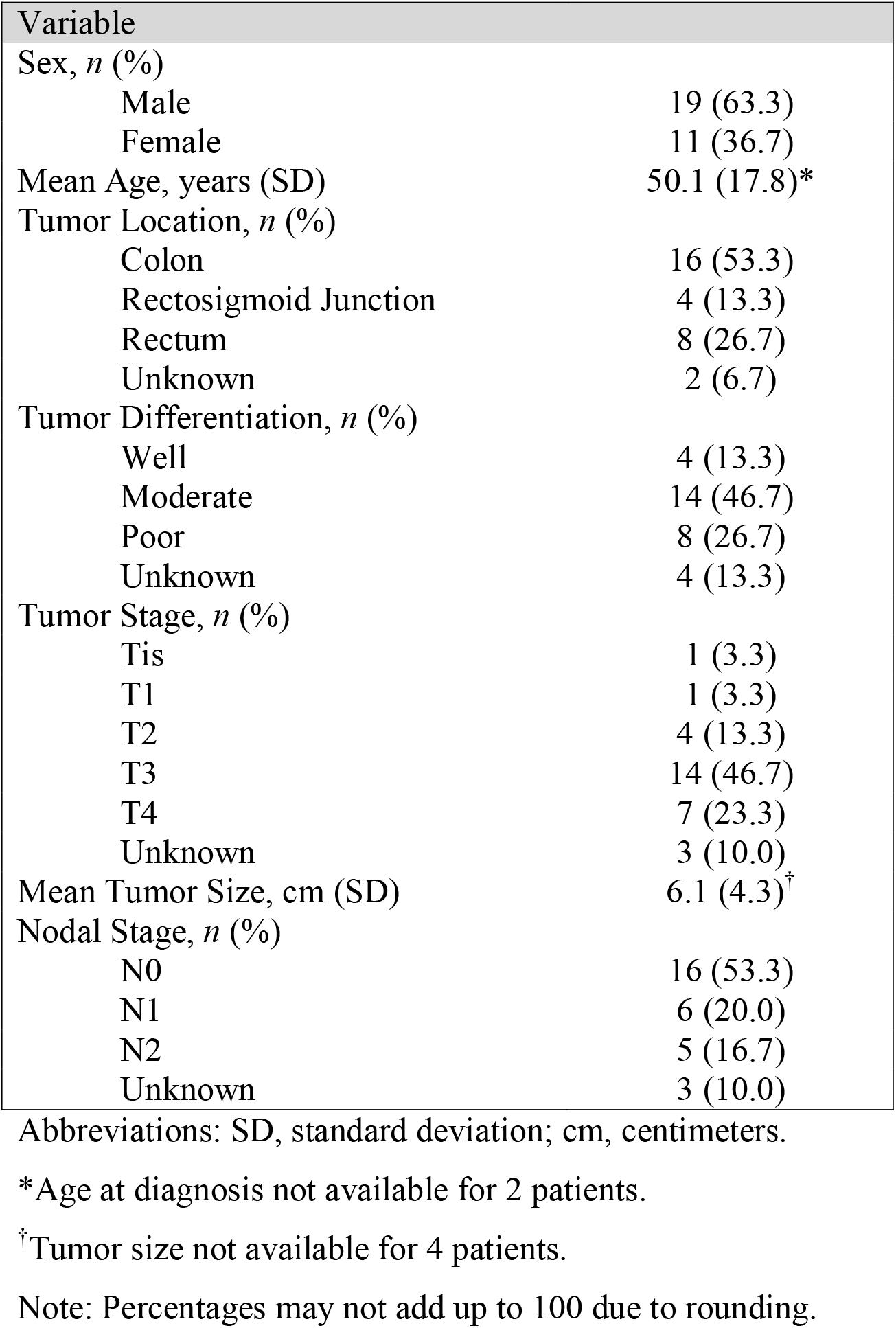
Patient and tumor characteristics (*n* = 30).

| Variable |  |
| --- | --- |
| Sex, *n* (%) |  |
| Male | 19 (63.3) |
| Female | 11 (36.7) |
| Mean Age, years (SD) | 50.1 (17.8)* |
| Tumor Location, *n* (%) |  |
| Colon | 16 (53.3) |
| Rectosigmoid Junction | 4 (13.3) |
| Rectum | 8 (26.7) |
| Unknown | 2 (6.7) |
| Tumor Differentiation, *n* (%) |  |
| Well | 4 (13.3) |
| Moderate | 14 (46.7) |
| Poor | 8 (26.7) |
| Unknown | 4 (13.3) |
| Tumor Stage, *n* (%) |  |
| Tis | 1 (3.3) |
| T1 | 1 (3.3) |
| T2 | 4 (13.3) |
| T3 | 14 (46.7) |
| T4 | 7 (23.3) |
| Unknown | 3 (10.0) |
| Mean Tumor Size, cm (SD) | 6.1 (4.3)<sup>†</sup> |
| Nodal Stage, *n* (%) |  |
| N0 | 16 (53.3) |
| N1 | 6 (20.0) |
| N2 | 5 (16.7) |
| Unknown | 3 (10.0) |

Abbreviations: SD, standard deviation; cm, centimeters.
\*Age at diagnosis not available for 2 patients.
<sup>†</sup>Tumor size not available for 4 patients.
Note: Percentages may not add up to 100 due to rounding.

### Sample and sequencing metrics

An average of 265 ng (SD, 290 ng) was extracted from each FFPE tumor sample. A mean of 5.9 ng (SD, 7.1 ng) of DNA was used as input for whole genome library preparation. Shallow WGS generated an average of 27.9 million reads (SD, 24.8 million), with a mean unique depth of 0.22× (SD, 0.21×). Based on ichorCNA results, median tumor cellularity was 13.6% (IQR, 0.3% to 26.9%).

### Copy number aberrations

Two representative ichorCNA plots showing the copy number alterations across the genome are shown in Figure 1A. Patient CRC-06 is a male in the fifth decade of life with a well-differentiated, 7.5 cm tumor of the colon with concomitant nodal disease. Patient CRC-25 is a male in the second decade of life with poorly differentiated rectal adenocarcinoma *in situ* (Tis). Both tumors exhibit copy number gains in chromosomes 8 and 19, as well as copy number losses in chromosome 18. The genome-wide copy number alteration profiles of tumors for which sample cellularity is estimated to be ≥ 3% by ichorCNA are shown in Figure 1B (*n* = 25).

**Figure 1:**
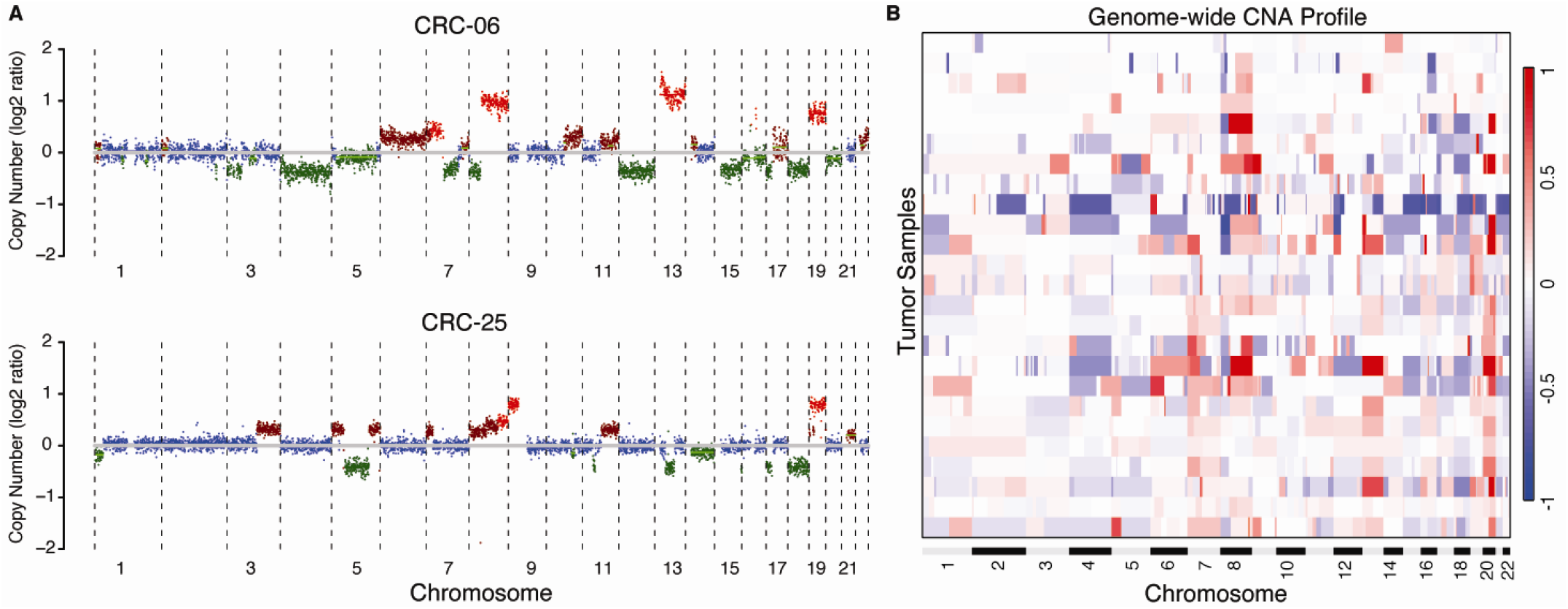
**(A)** Representative ichorCNA plots showing genome-wide copy number alterations in each 500 kb window. **(B)** Genome-wide copy number alteration profiles for samples with tumor cellularity ≥ 3% as estimated by ichorCNA (*n* = 25).

Using GISTIC2 across the 25 samples with tumor cellularity ≥ 3%, we identified two genomic regions with copy number gains exceeding the threshold for a statistically significant recurrent alteration: 8q24.22 and 13q12.3 (Figure 2). Meanwhile, region 8p21.3 demonstrated copy number loss exceeding similar cohort-wide thresholds for recurrence (Figure 2).

**Figure 2:**
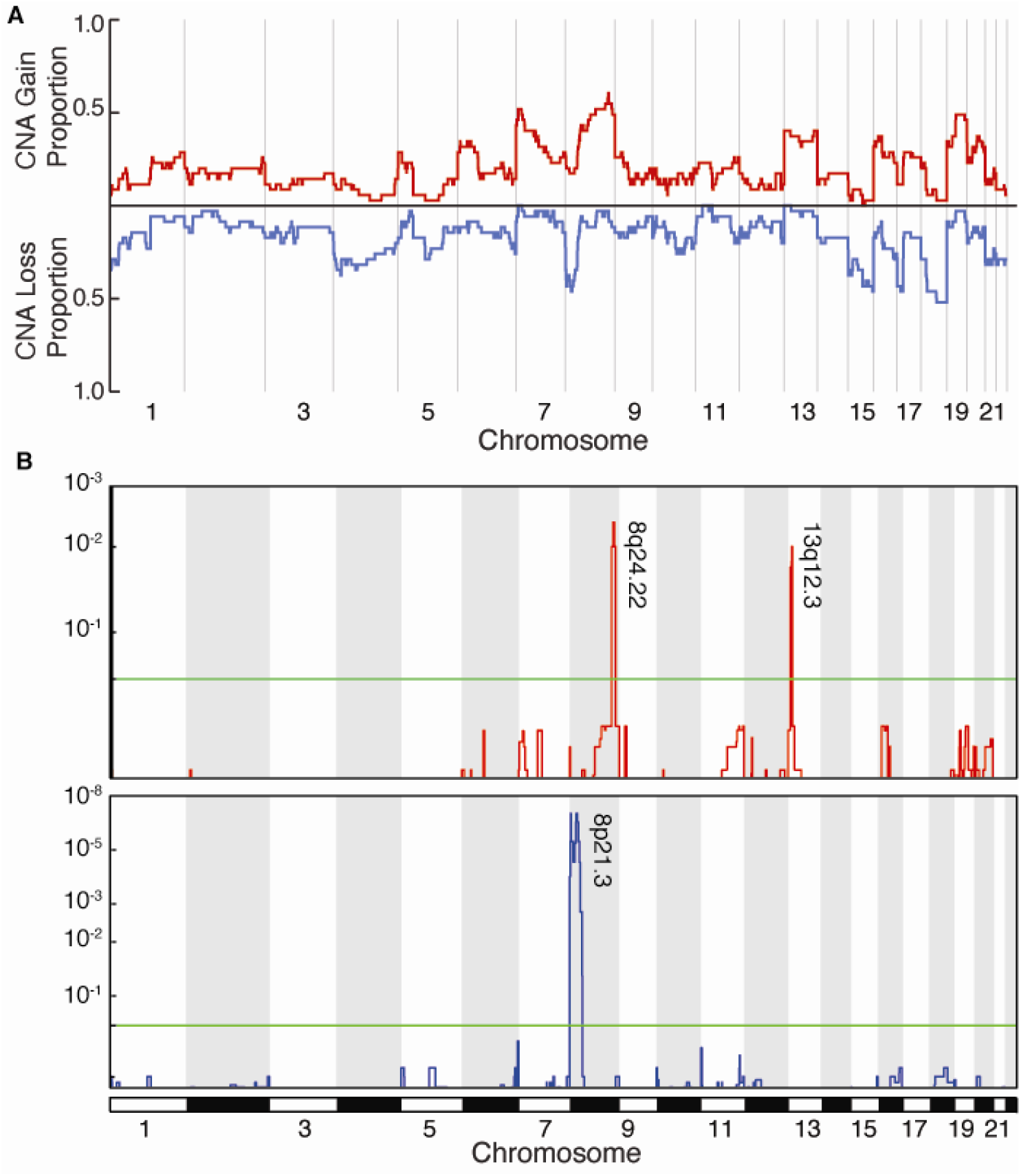
**(A)** Proportional copy-number gains and losses by chromosome across samples with tumor cellularity ≥ 3%. **(B)** Focal amplifications (top panel) and deletions (bottom panel) across samples with tumor cellularity ≥ 3%. The y-axis represents the GISTIC2.0 *q*-value for each locus. The horizontal green line represents the significance threshold (*q*-value of 0.25).^12^

## Discussion

In this new collaboration between academic institutions in Pakistan and the United States, we were able to successfully analyze copy number alterations in a cohort of 30 Pakistani patients with CRC using shallow WGS. We found recurrent copy number alterations in three genomic regions: copy number gains in 8q and 12q, and copy number loss in 8p. Specifically, 8q24.22 lies adjacent to the coding region for the *MYC* oncogene, overexpression of which is highly implicated in the pathogenesis of CRC through the *Wnt* signaling pathway.^14,15^ Region 8q24 is also notable as the location of *PVT1* and several other long non-coding RNA sequences implicated in many malignancies, including CRC.^16-18^ Region 13q12.3 is the location of *HMGB1*, another gene recently surmised to influence CRC pathogenesis.^19^ This region lies adjacent to the coding region of *CDX2*, which has been described as a lineage-survival oncogene when amplified.^20^ Chromosomal amplifications in 8q and 13q have been implicated in the adenoma to carcinoma sequence of CRC for the past several decades using hybridization techniques.^21-24^ Copy number losses in 8p have also been previously reported in prior array hybridization and sequencing-based studies of CRC.^22,24,25^ Gains in 8q and losses in 8p have also both been shown to be associated with lymph node positive disease and metastasis.^26-29^ Additional common copy number alterations seen in CRC such as gains in 20q and losses in 18q were observed in a proportion of patients, but they did not reach the threshold for statistically significant enrichment in this small cohort.^27,30^

We were able to identify several challenges that will need to be addressed to ensure the success of future efforts to analyze of cancer genomes in native South Asian populations. One notable challenge was the extent of degradation of nucleic acids in FFPE tissue, particularly for samples stored for longer periods. We and others have described whole genome and whole exome sequencing from FFPE DNA samples in the past.^31^ In addition, several commercial service providers routinely profile tumor DNA from FFPE blocks to identify actionable cancer mutations, and most centers in the United States have switched to nucleic acid-friendly fixation protocols.^32^ In the current study, 13 samples initially identified from the pathology archives were deemed ineligible for analysis because the DNA was too degraded or not amplifiable, and sequencing libraries could not be prepared. This experience demonstrates a need to evaluate and improve fixation protocols in LMICs to ensure readiness for future genomic analyses. While DNA fragmentation in FFPE affected overall sample size of this study, the choice of shallow WGS as the analytical assay helped overcome some of these limitations, as it can be performed with limited amounts of input (in contrast to targeted sequencing).^33^ However, shallow WGS limited our ability to perform deeper characterization of single nucleotide variants in CRC driver genes such as *APC, KRAS, TP53*, or *PIK3CA*. This pilot study also does not have adequate power to find other known variants or discover additional drivers that may be unique to this population. Nevertheless, the approach outlined in the study provides the advantage of providing sufficient data for copy number alteration analysis with minimal DNA input and fewer than 30 million reads per sample.

Further investments are needed to address not only the lack of ethnic and geographic diversity of genomic atlases, but also to make precision medicine breakthroughs more accessible to LMICs. Since 2021, a multi-institutional effort in India known as the Indian Cancer Genome Atlas (ICGA) aims to build a cancer genomic database in the style of TCGA.^34^ Leaders of this initiative acknowledge the need for transnational collaboration with neighboring states in order to build a genomic network that is mutually beneficial for the entire subcontinent. Beyond regional working groups, global health partnerships between high-income countries (HICs) and LMICs can address local needs while mobilizing genomics infrastructure and personnel expertise in HICs. The resulting HIC-LMIC relationship promotes the development of local capacity and skill sets that improve global health equity. This study highlights the feasibility of such a collaboration that navigates long-distance sample shipping and coordination, material transfer agreements, and coordinated data acquisition.

In conclusion, this study demonstrates a successful HIC-LMIC collaboration that can expand our understanding of cancer genomics in South Asia. We highlight the utility of fast, reliable, and affordable shallow WGS for this purpose, which requires only a small amount of tumor DNA. In ongoing studies, we are expanding this global, interinstitutional collaboration to include prospectively collected tumor and blood samples. Our goal is that these studies can be harnessed to bridge the knowledge gap in cancer genomics, to build local capacity and technical skill, and to help expand the benefit of precision cancer medicine to patients in LMICs.

## Data Availability

All data produced in the present study are available upon reasonable request to the authors.

## References

1. Ferlay J EM, Lam F, Colombet M, Mery L, Piñeros M, Znaor A, Soerjomataram I, Bray F. Global Cancer Observatory: Cancer Today. International Agency for Research on Cancer. Accessed April 10, 2023. https://gco.iarc.fr/today

2. GLOBOCAN. Cancer Today. GLOBOCAN. https://gco.iarc.fr/

3. Bishehsari F, Mahdavinia M, Vacca M, Malekzadeh R, Mariani-Costantini R. Epidemiological transition of colorectal cancer in developing countries: environmental factors, molecular pathways, and opportunities for prevention. World J Gastroenterol. May 28 2014;20(20):6055–72. doi:10.3748/wjg.v20.i20.6055

4. Cancer Genome Atlas N. Comprehensive molecular characterization of human colon and rectal cancer. Nature. Jul 18 2012;487(7407):330–7. doi:10.1038/nature11252

5. Lee KS, Kwak Y, Nam KH, et al. c-MYC Copy-Number Gain Is an Independent Prognostic Factor in Patients with Colorectal Cancer. PLoS One. 2015;10(10):e0139727. doi:10.1371/journal.pone.0139727

6. Carrot-Zhang J, Chambwe N, Damrauer JS, et al. Comprehensive Analysis of Genetic Ancestry and Its Molecular Correlates in Cancer. Cancer Cell. May 11 2020;37(5):639–654 e6. doi:10.1016/j.ccell.2020.04.012

7. Li H, Durbin R. Fast and accurate short read alignment with Burrows-Wheeler transform. Bioinformatics. Jul 15 2009;25(14):1754–60. doi:10.1093/bioinformatics/btp324

8. Li H. Aligning sequence reads, clone sequences and assembly contigs with BWA-MEM. arXiv pre-print server. 2013-05-26 2013;doi:None arxiv:1303.3997

9. Li H, Handsaker B, Wysoker A, et al. The Sequence Alignment/Map format and SAMtools. Bioinformatics. Aug 15 2009;25(16):2078–9. doi:10.1093/bioinformatics/btp352

10. Adalsteinsson VA, Ha G, Freeman SS, et al. Scalable whole-exome sequencing of cell-free DNA reveals high concordance with metastatic tumors. Nat Commun. Nov 6 2017;8(1):1324. doi:10.1038/s41467-017-00965-y

11. Quinlan AR, Hall IM. BEDTools: a flexible suite of utilities for comparing genomic features. Bioinformatics. Mar 15 2010;26(6):841–2. doi:10.1093/bioinformatics/btq033

12. Mermel CH, Schumacher SE, Hill B, Meyerson ML, Beroukhim R, Getz G. GISTIC2.0 facilitates sensitive and confident localization of the targets of focal somatic copy-number alteration in human cancers. Genome Biol. Apr 28 2011;12(4):R41. doi:10.1186/gb-2011-12-4-r41

13. Bezanson J, Edelman A, Karpinski S, Shah VB. Julia: A Fresh Approach to Numerical Computing. SIAM Review. 2017;59(1):65–98. doi:10.1137/141000671

14. He T-C, Sparks AB, Rago C, et al. Identification of c-MYC as a Target of the APC Pathway. Science. 1998;281(5382):1509–1512. doi:10.1126/science.281.5382.1509

15. Sansom OJ, Meniel VS, Muncan V, et al. Myc deletion rescues Apc deficiency in the small intestine. Nature. Apr 5 2007;446(7136):676–9. doi:10.1038/nature05674

16. Ozawa T, Matsuyama T, Toiyama Y, et al. CCAT1 and CCAT2 long noncoding RNAs, located within the 8q.24.21 ‘gene desert’, serve as important prognostic biomarkers in colorectal cancer. Ann Oncol. Aug 1 2017;28(8):1882–1888. doi:10.1093/annonc/mdx248

17. He F, Song Z, Chen H, et al. Long noncoding RNA PVT1-214 promotes proliferation and invasion of colorectal cancer by stabilizing Lin28 and interacting with miR-128. Oncogene. Jan 2019;38(2):164–179. doi:10.1038/s41388-018-0432-8

18. Gagliardi A, Francescato G, Ferrero G, et al. The 8q24 region hosts miRNAs altered in biospecimens of colorectal and bladder cancer patients. Cancer Med. Mar 2023;12(5):5859–5873. doi:10.1002/cam4.5375

19. Porter RJ, Murray GI, Hapca S, et al. Subcellular Epithelial HMGB1 Expression Is Associated with Colorectal Neoplastic Progression, Male Sex, Mismatch Repair Protein Expression, Lymph Node Positivity, and an ‘Immune Cold’ Phenotype Associated with Poor Survival. Cancers (Basel). Mar 20 2023;15(6)doi:10.3390/cancers15061865

20. Salari K, Spulak ME, Cuff J, et al. CDX2 is an amplified lineage-survival oncogene in colorectal cancer. Proc Natl Acad Sci U S A. Nov 13 2012;109(46):E3196–205. doi:10.1073/pnas.1206004109

21. Reichmann A, Martin P, Levin B. Chromosomal banding patterns in human large bowel cancer. Int J Cancer. Oct 15 1981;28(4):431–440. doi:10.1002/ijc.2910280407

22. Ried T, Knutzen R, Steinbeck R, et al. Comparative genomic hybridization reveals a specific pattern of chromosomal gains and losses during the genesis of colorectal tumors. Genes Chromosomes Cancer. 1996;15(4):234–245. doi:10.1002/(sici)1098-2264(199604)15:4<234::Aid-gcc5>3.0.Co;2-2

23. Meijer GA, Hermsen MA, Baak JP, et al. Progression from colorectal adenoma to carcinoma is associated with non-random chromosomal gains as detected by comparative genomic hybridisation. J Clin Pathol. 1998;51(12):901. doi:10.1136/jcp.51.12.901

24. Nakao K, Mehta KR, Fridlyand J, et al. High-resolution analysis of DNA copy number alterations in colorectal cancer by array-based comparative genomic hybridization. Carcinogenesis. Aug 2004;25(8):1345–57. doi:10.1093/carcin/bgh134

25. Xie T, G Da, Lamb JR, et al. A comprehensive characterization of genome-wide copy number aberrations in colorectal cancer reveals novel oncogenes and patterns of alterations. PLoS One. 2012;7(7):e42001. doi:10.1371/journal.pone.0042001

26. Ghadimi BM, Grade M, Liersch T, et al. Gain of Chromosome 8q23–24 Is a Predictive Marker for Lymph Node Positivity in Colorectal Cancer. Clin Cancer Res. 2003;9(5):1808–1814.

27. Golas MM, Gunawan B, Cakir M, et al. Evolutionary patterns of chromosomal instability and mismatch repair deficiency in proximal and distal colorectal cancer. Colorectal Dis. Feb 2022;24(2):157–176. doi:10.1111/codi.15946

28. Hermsen M, Postma C, Baak J, et al. Colorectal adenoma to carcinoma progression follows multiple pathways of chromosomal instability. Gastroenterology. Oct 2002;123(4):1109–19. doi:10.1053/gast.2002.36051

29. Joosse SA, Souche FR, Babayan A, et al. Chromosomal Aberrations Associated with Sequential Steps of the Metastatic Cascade in Colorectal Cancer Patients. Clin Chem. Oct 2018;64(10):1505–1512. doi:10.1373/clinchem.2018.289819

30. Voutsadakis IA. Chromosome 20q11.21 Amplifications in Colorectal Cancer. Cancer Genomics Proteomics. May-Jun 2021;18(3 Suppl):487–496. doi:10.21873/cgp.20274

31. Chehrazi-Raffle A, Muddasani R, Dizman N, et al. Ultrasensitive Circulating Tumor DNA Pilot Study Distinguishes Complete Response and Partial Response With Immunotherapy in Patients With Metastatic Renal Cell Carcinoma. JCO Precis Oncol. Apr 2023;7:e2200543. doi:10.1200/PO.22.00543

32. Milbury CA, Creeden J, Yip WK, et al. Clinical and analytical validation of FoundationOne(R)CDx, a comprehensive genomic profiling assay for solid tumors. PLoS One. 2022;17(3):e0264138. doi:10.1371/journal.pone.0264138

33. Chin SF, Santonja A, Grzelak M, et al. Shallow whole genome sequencing for robust copy number profiling of formalin-fixed paraffin-embedded breast cancers. Exp Mol Pathol. Jun 2018;104(3):161–169. doi:10.1016/j.yexmp.2018.03.006

34. Joshi S, Mishra R, Kulkarni M, et al. Proceedings of the 3rd Indian Cancer Genome Atlas Conference 2022: Biobanking to Omics: Collecting the Global Experience. JCO Glob Oncol. Jan 2023;9:e2200176. doi:10.1200/GO.22.00176

